# COVID-19 in Children with Down Syndrome: Data from the Trisomy 21 Research Society Survey

**DOI:** 10.1101/2021.06.25.21259525

**Authors:** David Emes, Anke Hüls, Nicole Baumer, Mara Dierssen, Shiela Puri, Lauren Russel, Stephanie L. Sherman, Andre Strydom, Stefania Bargagna, Ana Cláudia Brandão, Alberto C.S. Costa, Brian Allen Chicoine, Sujay Ghosh, Anne-Sophie Rebillat, Giuseppina Sgandurra, Diletta Valentini, Tilman R. Rohrer, Johannes Levin, Monica Lakhanpaul, The Trisomy 21 Research Society COVID-19 Initiative

## Abstract

**Importance:** Adults with Down syndrome (DS) are at higher risk for severe outcomes of coronavirus disease 2019 (COVID-19), but further evidence is required to determine the exact risks for children with DS. The clinical features and epidemiological characteristics of COVID-19 in children with DS, and risk factors for severe outcomes, must be established to inform COVID-19 shielding advice and vaccination priority.

**Objective:** To determine risk factors for a severe course of COVID-19 in pediatric DS patients and to compare the prevalence of severe COVID-19 between pediatric patients with and without DS.

**Design:** This retrospective cohort study included pediatric cases (aged <18 years) with DS from the Trisomy 21 Research Society international survey and pediatric cases from the general population published by the US Centers for Disease Control and Prevention (COVID-NET) collected during the first wave of the COVID-19 pandemic (controls).

**Setting:** Cohorts included 328 children with DS (127 hospitalized, 39%) and 224 children without DS (all hospitalized) with COVID-19. Of the pediatric DS patients, 64.1% were from low-to-middle-income countries (LMICs), and 35.9% from high-income countries (HICs).

**Participants:** Clinicians, family members, or caregivers completed the survey on behalf of children with DS affected by COVID-19.

**Results:** Among the 328 COVID-19 patients with DS; older age, obesity, and epilepsy were significant risk factors for hospitalization; and age and thyroid disorder were significant risk factors for acute respiratory distress syndrome. The 127 hospitalized COVID-19 patients with DS had a higher incidence of cough, fever, nasal signs and shortness of breath than controls. Compared with controls, hospitalized children with DS (especially those from LMICs) had a higher prevalence of COVID-19-related medical complications (pneumonia, ARDS, acute renal failure).

**Conclusions and relevance:** Children with DS are at higher risk for severe COVID-19 than the general pediatric population. Efforts should be made to monitor the health of children and young people with DS during the ongoing pandemic and to report any COVID-19 signs and symptoms in a timely manner, especially for those who have comorbidities which are risk factors for severe COVID-19. When vaccination rollout for pediatric populations begins, children with DS should be prioritised.

**Key Points:** *Question:* What are the epidemiological and clinical characteristics of coronavirus disease 2019 (COVID-19) in paediatric patients with Down syndrome (DS)?

*Findings:* Hospitalised COVID-19 patients <18 years of age with DS from a range of countries had a higher incidence of respiratory symptoms, fever, and several medical complications from COVID-19 than patients without DS <18 years from the United States (US). Older age, obesity, and epilepsy were significant risk factors for hospitalisation among paediatric COVID-19 patients with DS; and age and thyroid disorder were significant risk factors for acute respiratory distress syndrome. Mortality rates were low in all paediatric COVID-19 patients (with and without DS), in contrast to previous findings in adults with DS (who exhibit higher mortality than those without DS).

*Significance:* Children with DS are at increased risk for more severe presentations of COVID-19. Efforts should be made to ensure comprehensive and early detection of COVID-19 in this population, and to identify children with DS who present comorbidities that pose a risk for a severe course of COVID-19. Children with DS should be prioritised for COVID-19 vaccination as part of children’s vaccination programmes.

## 1. Introduction

Down Syndrome (DS) is associated with genetic factors and medical comorbidities that may lead to increased vulnerability to a severe course of coronavirus disease 2019 (COVID-19), the illness caused by the severe acute respiratory syndrome coronavirus 2 (SARS-CoV-2). Immune dysregulation among individuals with DS increases vulnerability to viral infections, anatomical airway features increase the vulnerability to respiratory illnesses^1,2,3^. Respiratory illnesses are a major cause of premature mortality in people with DS^4,5^ and children with DS show a high susceptibility to recurrent respiratory infections^6^. Indeed, many non-respiratory comorbidities of DS, such as obesity, diabetes or heart conditions, are known risk factors for morbidity and mortality from COVID-19 in the general population^2,3^.

Unlike in the adult population, limited data are available on children with DS and COVID-19. Some reports have highlighted a few cases of children who had one or more comorbidities, including cardiovascular anomalies, obesity, and/or obstructive sleep apnoea^7,8^, but these reports were based on only 55 and 4 participants respectively.

This study aims to describe the epidemiological and clinical characteristics of COVID-19 in children with DS, including risk factors for severe course of illness, compared to those in the general paediatric population. In this study, severe COVID-19 is assessed holistically by comparing the rate of hospitalisation (including rate of mechanical ventilation and intensive care unit (ICU) admission), acute respiratory distress syndrome (ARDS) and shortness of breath, in addition to mortality. Due to limited data availability, control data (children with COVID-19 from the general population) were only available from the US. As our data on children with DS were drawn from countries with varying income levels, we explored the differential course of pediatric COVID-19 cases from low-to-middle-income countries (LMICs) and from high-income countries (HICs)^9^.

## 2. Methods

### T21RS DS survey

We used data from the Trisomy 21 Research Society (T21RS) international survey, the largest survey of individuals with DS who had COVID-19^2^. Caregivers and clinicians caring for people with DS provided information about the symptoms, outcomes and existing health conditions of COVID-19 patients using an anonymous online format. The clinician surveys also asked about COVID-related treatments and medical complications. From all pediatric cases under 18 years of age reported to T21RS from April 9, 2020 and October 22, 2020, after excluding patients with missing relevant information such as age, gender or clinical information, our sample included 328 paediatric COVID-19 patients with DS, who accounted for 31.4% of all COVID-19 patients with DS registered in the survey. We excluded duplicated participants identified based on age, gender and country and other specific demographics. Because this study relies on data from the first wave of the pandemic, 29% (95/328) of the paediatric cases reported in the T21RS dataset had not been tested for COVID-19 but were symptomatic (Table S4).

Each participating institution obtained IRB/ethics approval. The study was performed according to the Declaration of Helsinki and national guidelines and regulations for data privacy and all participants who completed the questionnaires provided informed consent. All data were anonymized according to good clinical practice guidelines and data protection regulations (Table S6).

### US COVID-NET Comparison Data

To contrast the COVID-19-related signs, symptoms, and medical complications with those observed in the general population, we compared the hospitalized T21RS DS cases to published results from paediatric cases reported by the COVID-19–Associated Hospitalization Surveillance Network (COVID-NET). COVID-NET is a US population-based surveillance system that collects data on laboratory-confirmed COVID-19–associated hospitalisations (Kim et al., 2020). During March 1–July 25, 2020, 576 hospitalized paediatric COVID-19 cases were reported to the COVID-NET^10^.

### Statistical analyses

We used descriptive statistics to show the demographic information, outcomes, COVID-19 symptoms and comorbidities of the participants included in our analyses. Categorical data were described as frequencies (percentages), and quantitative variables are provided as the mean (SD) or median (IQR). We used Fisher’s exact tests to compare characteristics of the COVID-NET controls to those observed in COVID-19 patients with DS. Due to limited data availability, we only had control data from the US (COVID-NET). To detect possible country-specific differences in the healthcare system and access to hospitals, we stratified the COVID-19 patients with DS from the T21RS survey by income level of the country of residence. Countries were categorized as HICs (USA, Canada, Western Europe, Argentina, Australia) or LMICs (Brazil, India, Iran, Egypt, Belarus, South Africa, Mexico, Costa Rica, Bangladesh, Nepal) based on the World Bank income classification system^11^.

To identify risk factors associated with adverse outcomes of COVID-19 in children with DS, we estimated associations between potential risk factors and admission to hospital, diagnosis of acute respiratory distress syndrome (ARDS), and presence of shortness of breath using logistic regression analyses. All association analyses were restricted to symptomatic COVID-19 cases from the T21RS survey data, controlling for data source (caregiver versus clinician survey), living situation, level of intellectual disability, age, gender, and country of residence.

All data analyses were done using R (version 4.0.0).

## 3. Results

This retrospective cohort study included 328 paediatric patients <18 years of age with DS and COVID-19. Of the individuals for whom we had relevant information, 38.8% (127/325) were hospitalised. Mean age was 9.55 (SD=5.24) years (8.47 (SD=4.99) for non-hospitalised, 11.37 (SD=5.07) for hospitalised). Of the patients for whom country information was available, 42.9% (137/319) were from India, 17.6% (56/319) were from Brazil, 11.3% (36/319) from the UK, 9.4% (30/319) from the USA, 3.8% (12/319) from Spain and 15% (48/319) from other countries (Table 1). Note that some information was missing for certain patients - percentages reflect the portion of individuals for whom data were available.The age distribution was slightly different between the T21RS individuals with Down syndrome and the CDC controls without Down syndrome (Table S7). The CDC controls involved more newborns <1 year of age (CDC controls: 27.3%, T21RS: 4.3%) and the proportion of 5-11 year-olds was higher among the study population with Down syndrome (CDC controls: 16.8%, T21RS: 36.3%). The gender distribution was comparable between both studies (CDC controls: 50.7% male, T21RS: 47.4% male).

**Table 1.** **T21RS** study characteristics grouped by hospital admission

|  | Overall | Not hospitalized | Hospitalized |
| --- | --- | --- | --- |
| n | 328 | 197 | 127 |
| Additional information through follow-up (%)<sup>#</sup> | 29 ( 8.8) | 25 (12.7) | 4 ( 3.1) |
| Country (%) |  |  |  |
| India | 137 (42.9) | 52 (27.1) | 85 ( 69.1) |
| Brazil | 56 (17.6) | 44 (22.9) | 11 ( 8.9) |
| United Kingdom | 36 (11.3) | 28 (14.6) | 8 (6.5) |
| United States | 30 ( 9.4) | 22 (11.5) | 7 (5.7) |
| Spain | 12 ( 3.8) | 9 ( 4.7) | 3 ( 2.4) |
| other | 48 (15.0) | 37 (19.3) | 9 ( 7.3) |
| Age (mean (SD)) | 9.55 (5.24) | 8.47 (4.99) | 11.37 (5.07) |
| Male (%) | 173 (52.7) | 97 (48.5) | 74 ( 58.3) |
| Ethnicity (%) |  |  |  |
| South Asian | 142 (43.2) | 54 (27.4) | 88 ( 69.3) |
| White | 110 (33.4) | 89 (44.2) | 21 ( 16.5) |
| Latin American | 31 ( 9.4) | 23 (11.7) | 7 ( 5.5) |
| Black | 6 ( 1.8) | 3 ( 1.5) | 3 ( 2.4) |
| Arab | 1 ( 0.3) | 1 ( 0.5) | 0 ( 0.0) |
| West Asian | 1 ( 0.3) | 1 ( 0.5) | 0 ( 0.0) |
| Admixed | 12 ( 3.6) | 9 ( 4.6) | 3 ( 2.4) |
| Unknown | 26 ( 7.9) | 19 ( 9.6) | 5 ( 3.9) |
| Living situation (%) |  |  |  |
| Living at home with family | 304 (98.1) | 190 (97.9) | 110 ( 98.2) |
| Living alone with support | 1 ( 0.3) | 0 ( 0.0) | 1 ( 0.9) |
| Small group home with support | 2 ( 0.6) | 2 ( 1.0) | 0 ( 0.0) |
| Residential care facility | 2 ( 0.6) | 1 ( 0.5) | 1 ( 0.9) |
| Other | 1 ( 0.3) | 1 ( 0.5) | 0 ( 0.0) |
| Type of trisomy 21 (%) |  |  |  |
| Full/standard | 274 (88.4) | 167 (87.9) | 105 ( 89.7) |
| Mosaic | 29 ( 9.4) | 17 ( 8.9) | 11 ( 9.4) |
| Partial trisomy | 2 ( 0.6) | 2 ( 1.0) | 0 ( 0.0) |
| Translocation | 5 ( 1.6) | 4 ( 2.1) | 1 ( 0.9) |
| Level of intellectual disability (%) |  |  |  |
| Borderline/normal/mild | 83 (30.0) | 59 (37.1) | 23 ( 20.2) |
| Moderate | 172 (62.1) | 91 (57.2) | 78 ( 68.4) |
| Severe/Profound | 22 ( 7.9) | 9 ( 5.7) | 13 ( 11.4) |
| Admitted to hospital (%) | 127 (39.2) | 0 ( 0.0) | 127 (100.0) |
| Days in hospital (mean (SD))* | 11.03 (5.18) | NaN** (NA**) | 11.03 (5.18) |
| Admitted to ICU (%)* | 48 (38.4) | 0 ( NaN) | 48 ( 38.4) |
| Days in ICU (mean (SD))* | 6.91 (2.52) | NaN (NA) | 6.91 (2.52) |
| Mechanical ventilation (%)* | 28 (16.1) | 0 ( 0.0) | 28 ( 24.8) |
| Clinical situation at last evaluation (%) |  |  |  |
| Currently in hospital with symptoms | 38 (12.5) | 0 ( 0.0) | 37 ( 31.4) |
| Died | 4 ( 1.3) | 0 ( 0.0) | 4 ( 3.4) |
| Not currently in hospital but with symptoms | 45 (14.8) | 34 (18.6) | 10 ( 8.5) |
| Other | 12 ( 3.9) | 9 ( 4.9) | 3 ( 2.5) |
| Recovered from COVID-19 | 197 (64.8) | 132 (71.1) | 64 ( 54.2) |
| Tested positive but still no symptoms | 8 ( 2.6) | 8 ( 4.4) | 0 ( 0.0) |

\*Only answered if admitted to hospital; #Information that was missing in the original report was imputed by information provided through a follow-up survey (clinician follow-up for the family survey and family follow-up for the clinician survey, see methods for more details); % were calculated after excluding missing information
\*\*Not Applicable

Hospitalised children with DS had significantly higher rates of cough (53.7% vs 29.5% in controls), fever (94.3% vs 54% in controls), nasal signs / congestion (65.0% vs 23.7% in controls) and shortness of breath (60.2% vs 22.3% in controls) than the control group (Table 2). Hospitalised children with DS did not experience a significantly different prevalence of abdominal symptoms in comparison to the control group.

**Table 2.**
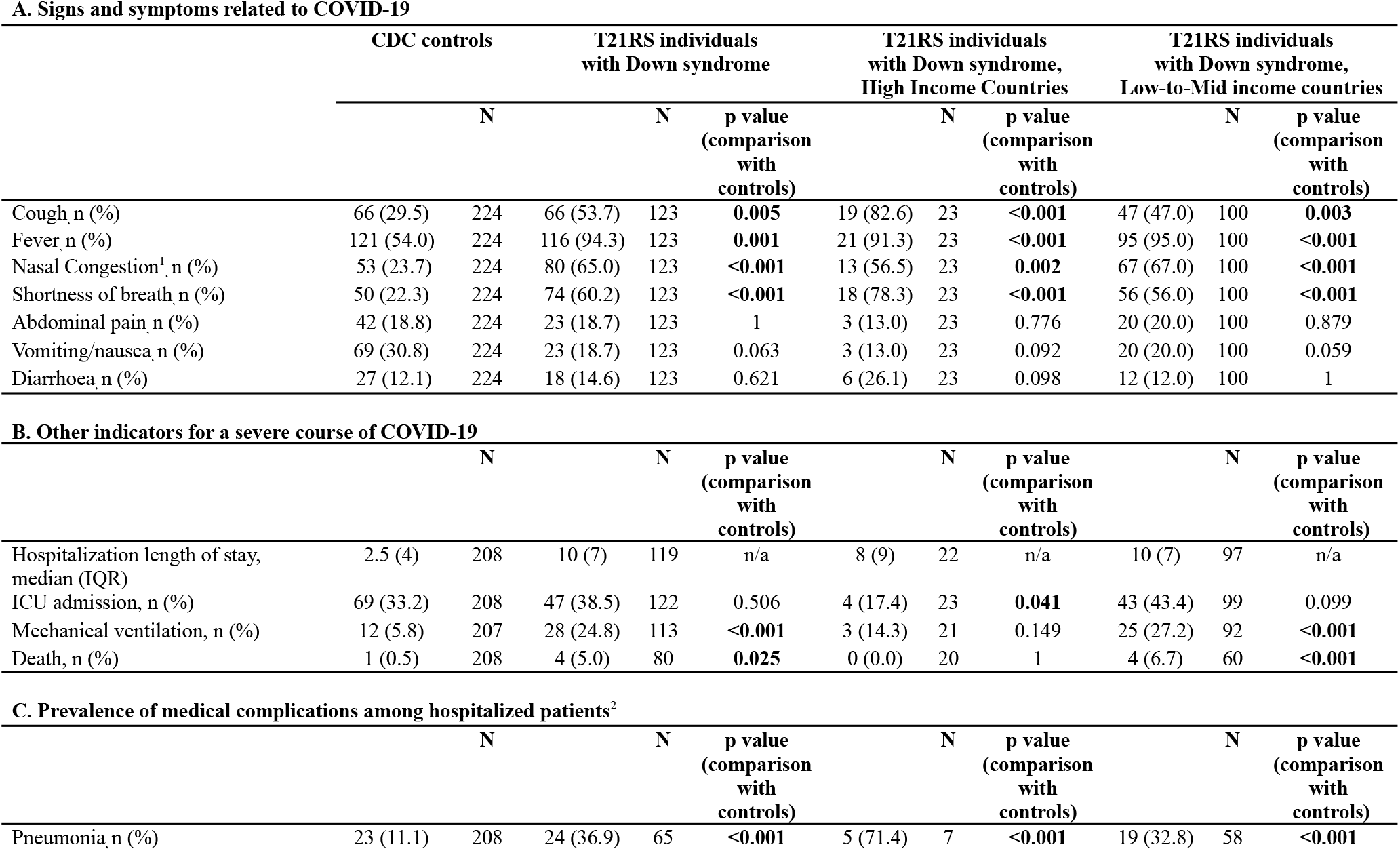
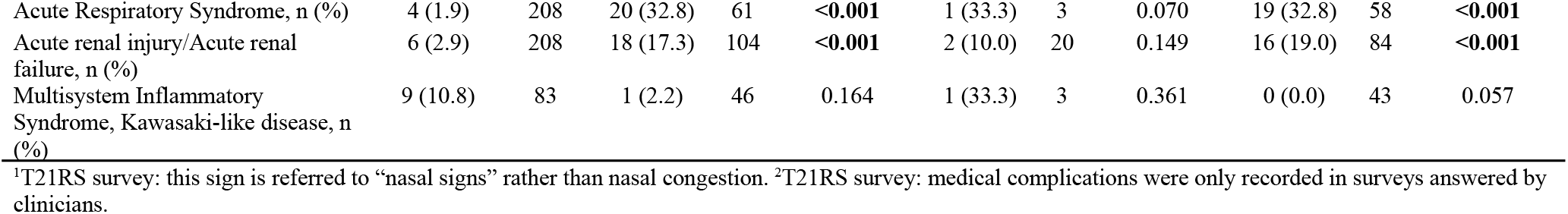
Signs and symptoms related to COVID-19 in hospitalized individuals with and without Down syndrome stratified by High Income and Low-to-Middle Income Countries.

Medical complications were more prevalent among children with DS than among controls. Children with DS had significantly higher rates of pneumonia (36.9% vs. 11.1% controls), ARDS (32.8% DS vs. 1.9% controls) and acute renal failure (17.3% DS vs. 2.9% controls). There was no significant difference in the prevalence of Multisystem Inflammatory Syndrome between COVID-19 patients with and without DS.

Children with DS had similar rates of ICU admission (38.4% DS vs 33.2% controls), but were more often put on mechanical ventilation (24.8% DS vs 5.8% controls). Hospitalized patients with DS had a higher mortality rate compared with controls (5.0% DS vs 0.5% controls) but all deaths occurred among children from LMIC.

Next, we examined potential risk factors for severe COVID-19 (shortness of breath, hospitalisation and ARDS) among COVID-19 patients with DS (Figure 2). Older age (OR 1.75 for 5 additional years [1.37-2.23]), obesity (OR 2.27 [1.12-4.59]) and epilepsy (OR 3.97 [1.65-9.58]) were all significant risk factors for hospitalisation among COVID-19 patients with DS; older age (OR 2.29 for 5 additional years [1.26-4.15]) and thyroid disorder (OR 3.40 [1.03-11.9]) were significant risk factors for ARDS; and older age (OR 1.31 for 5 additional years [1.05-1.63]) was a significant risk factor for shortness of breath.

**Figure 1.**
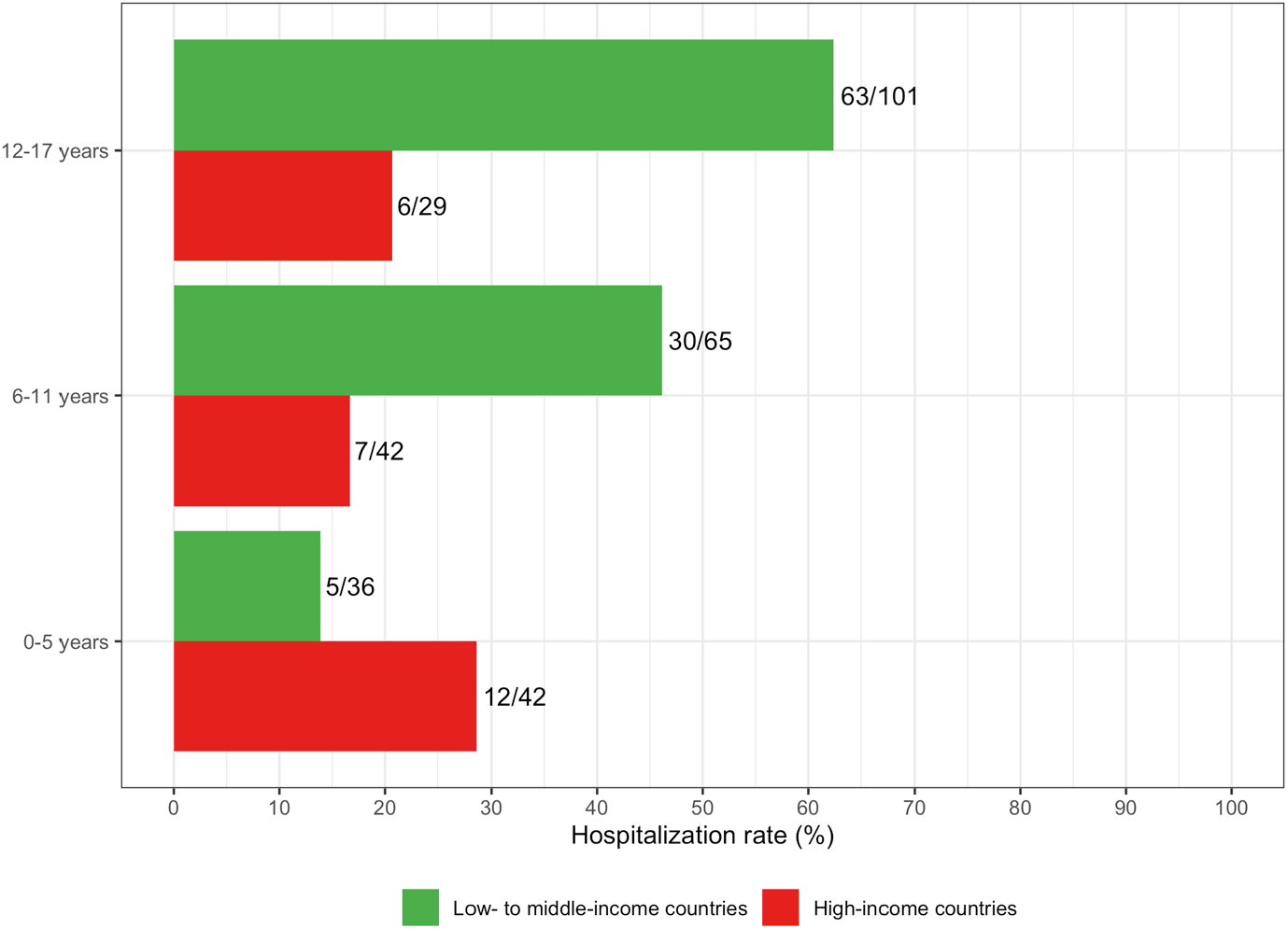
Hospitalization rate of children with Down syndrome and COVID-19 in low-to middle-income countries (Brazil, India, Iran, Egypt, Belarus, South Africa, Mexico, Costa Rica, Bangladesh, Nepal) and high-income countries (USA, Canada, Western Europe, Argentina, Australia)

**Figure 2.**
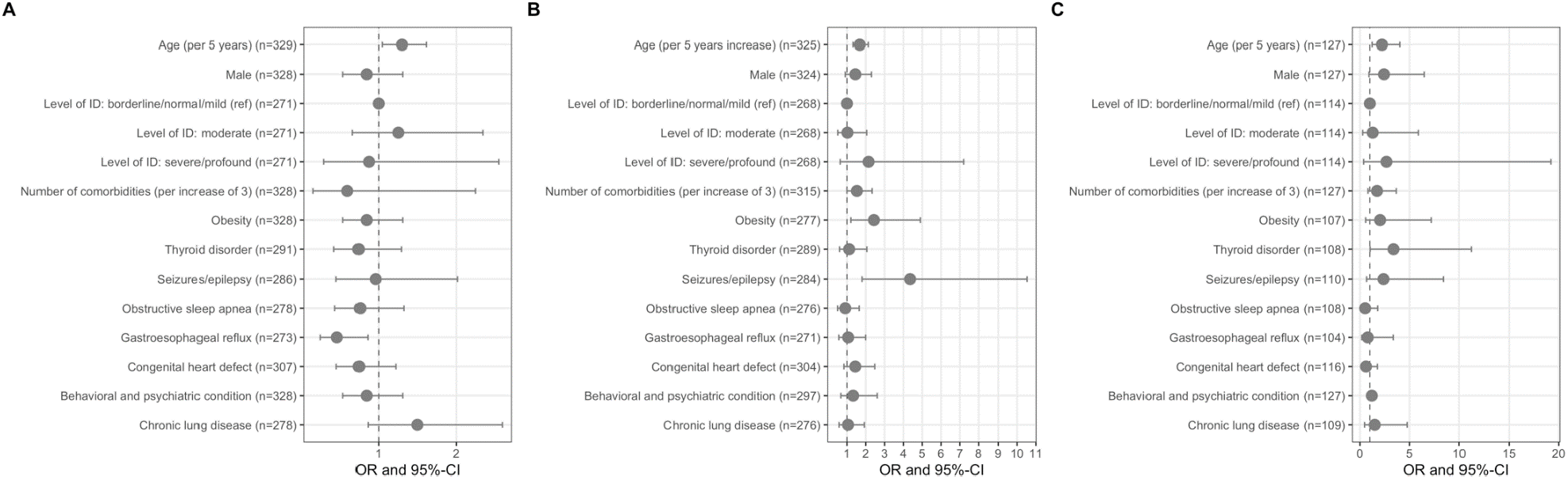
Risk factors associated with adverse outcomes of COVID-19 in children with Down syndrome. Associations with **A**. shortness of breath **B**. hospitalization and **C**. acute respiratory distress syndrome (ARDS) in symptomatic COVID-19 patients with Down syndrome from the T21RS survey estimated in adjusted logistic regression models (odds ratios (OR) and 95%-confidence intervals (95%-CI)). Associations with age and gender were adjusted for the data source (caregiver versus clinician survey) and associations with living situation, level of IDD and comorbidities were adjusted for age, gender, data source and country of residence. Abbreviations: IDD, intellectual and developmental disabilities; ref, reference category.

The pandemic has posed challenges to healthcare systems globally, and the nature of pandemic response may differ depending on country income status. We stratified our analysis by country income level to explore this. First, we examined hospitalisation rates by age group and country income status. Patients with DS from LMICs had a higher rate of hospitalisation (48.5%) than the sample as a whole (39.0%), except for among children under 5 years of age. In LMICs, hospitalisation rate increased with age group, consistent with our finding that older age was a risk factor for hospitalisation in our sample (Fig. 2). In HICs, the hospitalisation rate was highest for children under 5, but was similar for other age groups.

We compared the symptoms and signs of COVID-19 between the control group (all hospitalized) and hospitalised children with DS from the HICs and the LMICs. The prevalence of cough (82.6% in HICs, 46.5% in LMICs, 29.5% in controls), fever (91.3% in HICs, 95% in LMICs, 54% in controls), nasal congestion (56.5% in HICs, 66.7% in LMICs, 23.7% in controls) and shortness of breath (78.3% in HICs, 55.6% in LMICs, 22.4% in controls) was significantly higher among children with DS, independent of the country income status (although the latter was higher in HICs than in LMICs) (Table 2).

On average, children with DS spent more days in the hospital than children from the general population, independent of their country income status (controls: median of 2.5 days, HICs: median of 8 days, LMICs: median of 10 days). Prevalence of ICU admission was not different among children with DS from LMICs (43.4%), or HIC (17.4%), in comparison to controls (33.2%). The prevalence of mechanical ventilation was higher among children with DS from LMICs (27.2%) than among controls (5.8%) (p<0.001), but for children with DS from HICs (14.3%) there was no statistically significant difference from controls. Lastly, with respect to mortality rates, all deaths among hospitalized children with DS were from LMICs, leading to estimates of mortality of 6.7% in LMICs, with no deaths occurring in either controls or children with DS from HICs.

## 4. Discussion

Previous studies of individuals with DS suggest that, as in the general population, children do not face the same risk for COVID-19-related mortality as older adults. However, as severe COVID-19 cannot be assessed by mortality alone, we aimed to evaluate the rate of several indicators for severe COVID-19 (rate of hospitalisation, use of mechanical ventilation and ICU admittance, ARDS, shortness of breath, and mortality) among 328 children with DS and compare these to 224 children from the general population.

### 4.1 Hospitalisation and Outcomes

In our study, hospitalised children with DS had significantly higher rates of cough, fever, nasal signs and shortness of breath than the control group. This could be interpreted as a more severe COVID-19, as reflected by the following evidence of poor outcomes: 1) significantly higher rates of medical complications, including pneumonia, ARDS and acute renal failure and 2) need for mechanical ventilation (24.8% vs 5.8% in controls). Mortality rates for children with DS were much lower than for adults with DS^2^, as has also been reported in general for the pediatric population^12^. We could not examine risk factors for mortality because there were very few deaths among children with and without DS. Our results in children with DS are broadly in line with previous works on COVID-19 in adults with DS and with a literature which suggests that people with DS may be more susceptible to viral and bacterial infections in terms of symptoms, complications and a severe course of illness^4,5,7,13,14,15,16^.

Risk factors for a severe course of COVID-19 are well-documented for the adult population, but less so for the paediatric population^2,17,18^. In our sample, many of the documented risk factors for COVID-19 morbidity were found to be significant, including older age, epilepsy and obesity. However, several known risk factors for a severe course of COVID-19 were not found to be statistically significant, including male sex^19^ and chronic lung disease^2^. This may reflect a lack of statistical power, and underscores the need for further study.

Pre-existing epilepsy as a risk factor for infection or for severe COVID-19 outcomes has not been established for those without DS^20,21^. However, there is a consensus that management of COVID-19 in people with epilepsy compared with those without may be more complicated, as would be true for those with DS and epilepsy. Some examples of complications include drug-drug interactions^22^; use of certain epilepsy medications that may affect the immune system^21^; fever related to COVID-19 that may increase the risk of seizures and lead to increased hospitalization^20^; or increased need for health care or emergency care that may lead to higher exposure to the virus^20,21^. In those with and without DS, seizures could be a neurological consequence of COVID-19, as reviewed in Niazkar et al.^23^. However, further insights into the link between pre-existing epilepsy as a risk factor for COVID-19 outcomes in DS would benefit from further research. The same can be said of the risks associated with thyroid disorder, a common comorbidity of DS. It is possible that infection affected the thyroid mechanism of children with DS, but the mechanism cannot be determined from the data used in this study. However, both SARS and COVID-19 are known to be associated with thyroid abnormalities in adult patients. SARS patients have been found to have decreased serum T3, T4, and TSH levels^24^. Thyroid hormone dysfunction affects the clinical course of COVID-19 as it increases mortality in critical illnesses such as ARDS, a major complication of COVID-19^24^. ACE2 requires the transmembrane serine protease 2 (TMPRSS2) TMPRSS2 to enter cells. In humans, the TMPRSS2 enzyme is encoded by the TMPRSS2 gene located on chromosome 21 (21q22.3 (OMIM 602060)). TMPRSS2 in turn forms a complex with the ACE2 receptor, enabling the virus to penetrate the cell surface directly and efficiently^25^. ACE2 and TMPRSS2 are highly expressed in the thyroid gland^26^. Further investigation is needed to elucidate whether the impact of SARS-CoV-2 on thyroid function is one of several factors in the development of ARDS in people, particularly children, with DS and/or primarily due to overexpression of TMPRSS2 on chromosome 21 via a gene-dose mechanism.

Finally, results varied by country income status for some attributes. The rate of hospitalisation for children younger than 5 years of age with DS in HICs, and for the US controls, was particularly high as compared to LMICs. This may be a result of neonates being infected in hospitals immediately after birth^10^. Equally, higher availability of resources could allow children under five in HICs to be hospitalised for minor non-COVID-related symptoms or signs of illness in general, regardless of whether or not they have severe COVID-19. The high rate of hospitalisation in this age group in HICs (compared with children with DS from LMICs) might not, therefore, be indicative of a more severe course of illness.

### 4.2 Implications for Policy and Practice

Our results suggest that children with DS are more likely to develop severe COVID-19 than the general paediatric population. However, it is recognised that a child’s welfare also needs to be considered. Shielding children may protect them from being infected but it will also impact their access to education, interaction with peers and could have an adverse impact on their development and well being. While this is true for all children, it is particularly important for children with DS who are likely to be affected even more by social isolation and loss of their daily routine and therapies. These risks must therefore be balanced with those of protecting children from the infection itself. Parents, health professionals and policy makers may therefore wish to consider the risks of infection to an individual child taking account of their likely exposure to the infection, existing comorbidities and family circumstances.

While we found that the risk of severe COVID-19 increased with the presence of certain comorbidities for our population of interest, we also found that children with DS are, in general, at increased risk of severe outcomes from this disease. We therefore recommend, when vaccination is rolled out for paediatric populations, that children with DS be prioritised.

The health community have also yet to reach a conclusive understanding of the lifelong effects of COVID-19. Thus a mild course of the disease may have long-term implications, and children with DS who experience a less severe course of illness or who are not hospitalised may still require annual reviews to ensure that their condition has not deteriorated.

### 4.3 Limitations and Areas for Future Research

One notable limitation of our study is that control data were only available from hospitalised paediatric patients in the US, entailing a comparison of patients from LMICs to controls in HICs. Furthermore, the age distribution of the children with and without Down syndrome differed and as we had only access to the summary statistics, we could not adjust our association analyses for age. Subsequent international studies should explore comparison with controls from a range of countries when suitable data become available. In addition, most of our cases from LMICs were from India (137/202), making generalisation of findings to other LMICs difficult.

When reporting the ethnicity of patients, we used WHO categories, but the way in which ethnicity was reported varied among countries. We had limited data concerning about black, indigenous populations, Asian and other children representing minority ethnic groups in HICs, and may have overlooked risks unique to those groups.

More attention should also be paid to non-hospitalised and asymptomatic paediatric COVID-19 patients with DS, who could experience long-term effects of COVID-19. Thus, future research should aim to determine if the symptoms of non-hospitalised children with DS differ from those of the general paediatric population. Because our control data pertained to hospitalised patients only, this analysis lay beyond the scope of this investigation. Future research must also investigate the negative psychosocial and developmental impact of being home-bound during the COVID-19 pandemic on children with DS: emerging survey-based studies are already underway to document potential outcomes in terms of weight gain, regression, risk of osteoporosis from lack of exercise and so forth. Our understanding of COVID-19 will be further enhanced by studying asymptomatic cases: as such, it will be important to include data from asymptomatic paediatric patients with COVID-19 (and their caregivers and clinicians) in future surveys now that testing has become more widely available.

## 5. Conclusions

Hospitalised children with DS had a higher prevalence of several symptoms of COVID-19 (cough, fever, nasal signs, shortness of breath) than hospitalised US controls; a higher prevalence of medical complications from COVID-19 (pneumonia, ARDS, acute renal failure), particularly among children with DS from LMICs; and were more likely to be placed on mechanical ventilation. Older age, obesity, and epilepsy were significant risk factors for hospitalisation; older age and thyroid disorder were significant risk factors for ARDS; and older age was a significant risk factor for shortness of breath. Efforts should be made to monitor the health of children and young people with DS during the ongoing pandemic and to report any COVID-19 signs and symptoms in a timely manner, especially for those who have comorbidities which are risk factors for severe COVID-19. When vaccination is rolled out for paediatric populations, children with DS should be prioritised.

## Data Availability

Data used are drawn from international surveys, which are cited and made accessible to readers

## Supplement

**Table S1.** Risk factors associated with adverse outcomes of COVID-19 in children with Down syndrome. Associations with shortness of breath, hospitalization and acute respiratory distress syndrome (ARDS) in symptomatic COVID-19 patients with Down syndrome from the T21RS survey estimated in adjusted logistic regression models (odds ratios (OR) and 95%-confidence intervals (95%-CI)).

|  | Shortness of Breath OR,<br>(CI) | Hospitalized OR,<br>(CI) | ARDS OR, (CI) |
| --- | --- | --- | --- |
| Age (per 5 years) | 1.31 (1.05-1.63) | 1.75 (1.37-2.23) | 2.29 (1.26-4.15) |
| Male | 0.81 (0.52-1.27) | 1.45 (0.90-2.34) | 2.44 (0.91-6.55) |
| Level of ID: borderline/normal/mild<br>(ref) | 1 (1-1) | 1 (1-1) | 1 (1-1) |
| Level of ID: moderate | 1.20 (0.63-2.30) | 0.95 (0.46-1.93) | 1.40 (0.28-6.91) |
| Level of ID: severe/profound | 0.83 (0.28-2.42) | 1.94 (0.57-6.55) | 2.68 (0.36-19.9) |
| Number of comorbidities (per<br>increase of 3) | 0.53 (0.14-2.05) | 1.48 (0.96-2.28) | 1.82 (0.84-3.97) |
| Obesity | 0.81 (0.52-1.27) | 2.27 (1.12-4.59) | 2.14 (0.58-7.91) |
| Thyroid disorder | 0.74 (0.42-1.29) | 1.19 (0.64-2.22) | 3.40 (1.03-11.9) |
| Seizures/epilepsy | 0.92 (0.44-1.92) | 3.97 (1.65-9.58) | 2.10 (0.58-7.66) |
| Obstructive sleep apnea | 0.77 (0.44-1.36) | 0.96 (0.52-1.76) | 0.67 (0.19-2.40) |
| Gastroesophageal reflux | 0.43 (0.24-0.79) | 1.00 (0.53-1.87) | 0.78 (0.18-3.37) |
| Congenital Heart Defect | 0.72 (0.44-1.18) | 1.39 (0.81-2.42) | 0.52 (0.18-1.54) |
| Behavioral and psychiatric condition | 0.81 (0.52-1.27) | 1.29 (0.65-2.55) | 1.22 (0.94-1.58) |
| Chronic Lung Disease | 1.47 (0.85-2.56) | 1.02 (0.56-1.87) | 1.31 (0.40-4.24) |

**Table S2.** Comorbidities reported by the T21RS study.

|  | **Overall** | **Patients with Shortness**<br>**of Breath** | **Hospitalized**<br>**Patients** | **Patients with**<br>**ARDS** |
| --- | --- | --- | --- | --- |
| n | 328 | 259 | 127 | 20 |
| Obesity | 65 (23.0) | 37 (29.4) | 41 (38.7) | 9 (56.3) |
| Thyroid disorder | 93 (28.4) | 34 (26.0) | 34 (31.2) | 9 (56.3) |
| Seizures/epilepsy | 38 (13.1) | 19 (14.8) | 28 (25.5) | 6 (37.5) |
| Obstructive sleep apnea | 100 (35.5) | 45 (36.3) | 42 (39.3) | 4 (25.0) |
| Gastroesophageal reflux | 87 (31.4) | 35 (28.0) | 36 (34.3) | 3 (21.4) |
| Congenital Heart Defect | 149 (47.5) | 57 (42.9) | 61 (51.7) | 8 (42.1) |
| Behavioral and psychiatric<br>condition | 103 (33.6) | 52 (38.8) | 60 (52.6) | 9 (52.9) |
| Chronic Lung Disease | 103 (36.5) | 58 (46.0) | 48 (44.0) | 9 (52.9) |

**Table S3.**
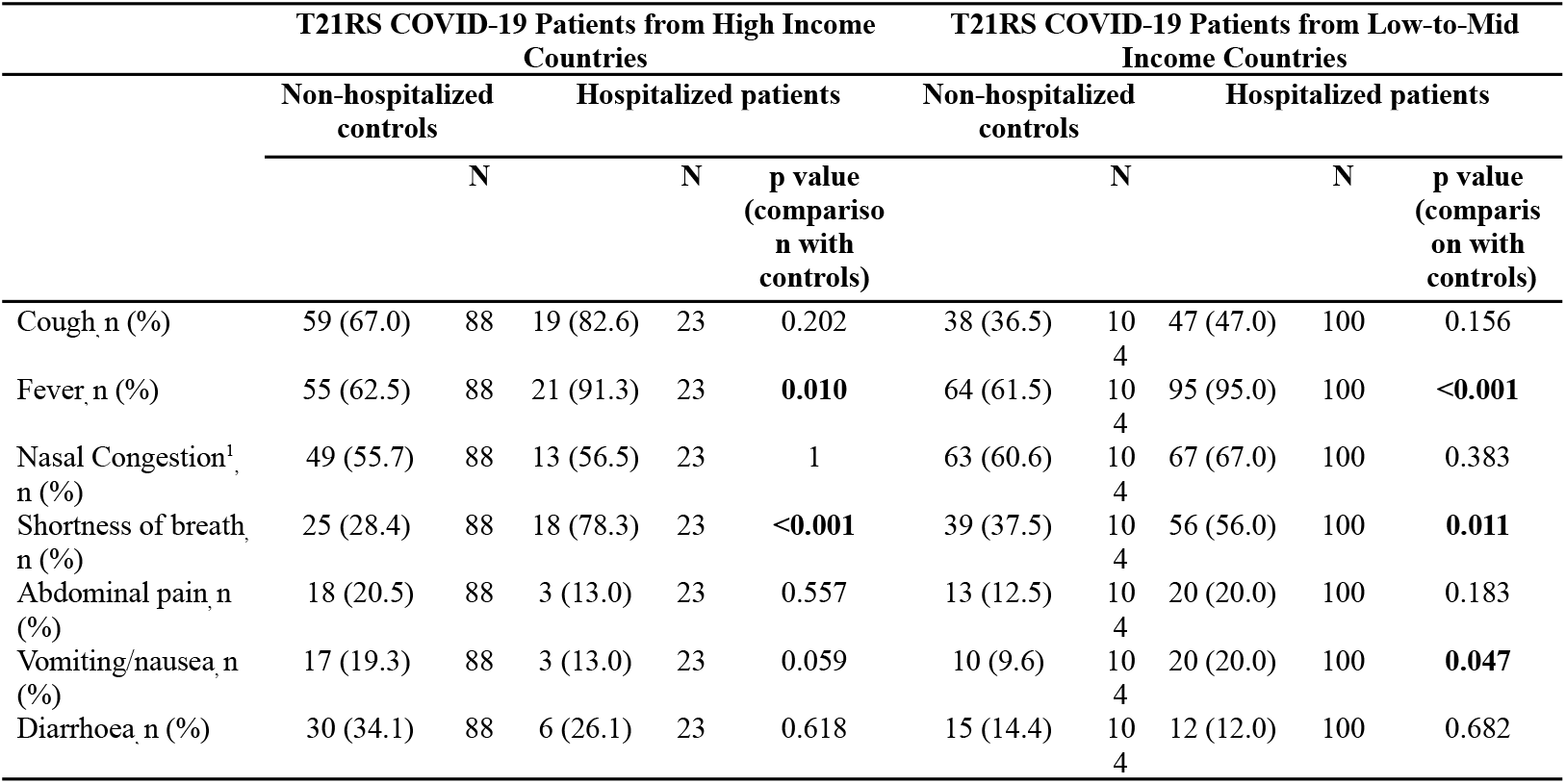
Signs and symptoms related to COVID-19 in individuals with Down syndrome comparing hospitalized with non-hospitalized patients, stratified by High Income and Low-to-Middle Income Countries.

**Table S4.** Proportion of the T21RS analysis sample with COVID-19 symptoms or positive test results.

|  | **Children with DS** |
| --- | --- |
| n | 328 |
| Signs and symptoms of COVID-19 (%) | 312 (95.4) |
| COVID-19 testing performed (%) | 233 (71.5) |
| Reason for COVID-10 testing (%) |  |
| Contact with affected individuals | 38 (16.6) |
| Routine testing | 3 (1.3) |
| Symptoms were present | 188 (82.1) |
| Results of the testing (%) |  |
| Negative | 16 (7.4) |
| Pending | 6 (2.8) |
| Positive | 194 (89.8) |

**Table S5.**
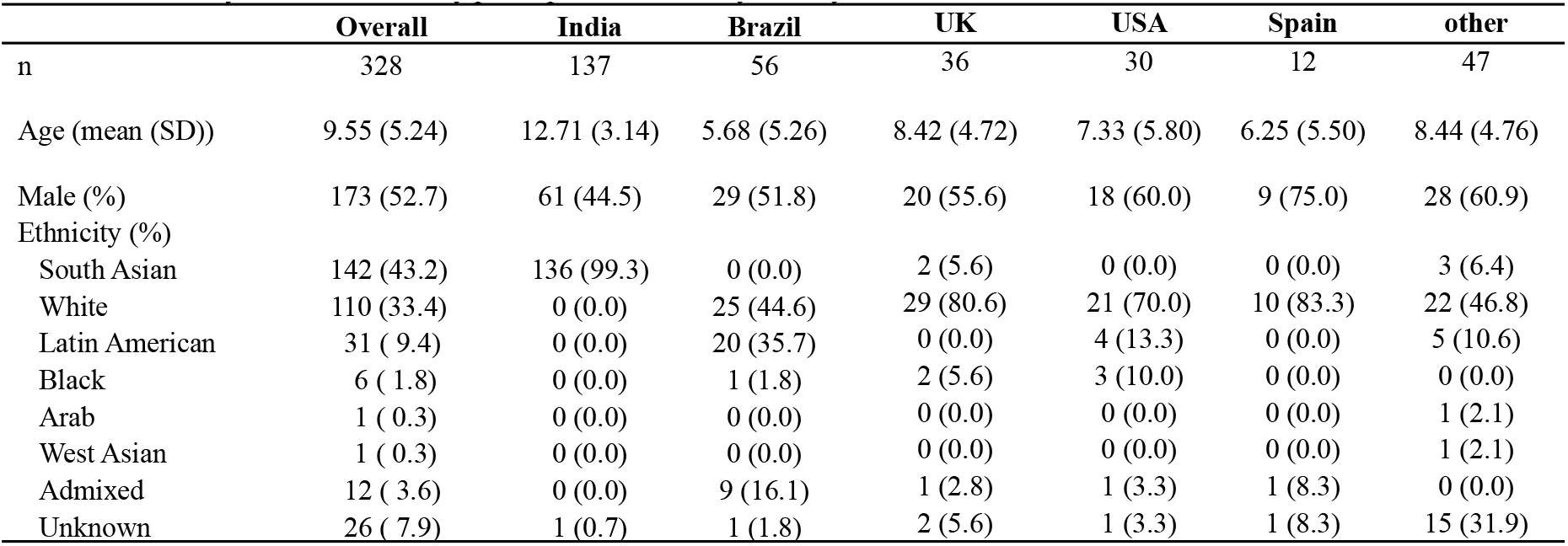
Ethnicity of the T21RS study participants stratified by country of residence.

|  | **Overall** | **India** | **Brazil** | **UK** | **USA** | **Spain** | **other** |
| --- | --- | --- | --- | --- | --- | --- | --- |
| n | 328 | 137 | 56 | 36 | 30 | 12 | 47 |
| Age (mean (SD)) | 9.55 (5.24) | 12.71 (3.14) | 5.68 (5.26) | 8.42 (4.72) | 7.33 (5.80) | 6.25 (5.50) | 8.44 (4.76) |
| Male (%) | 173 (52.7) | 61 (44.5) | 29 (51.8) | 20 (55.6) | 18 (60.0) | 9 (75.0) | 28 (60.9) |
| Ethnicity (%) |  |  |  |  |  |  |  |
| South Asian | 142 (43.2) | 136 (99.3) | 0 (0.0) | 2 (5.6) | 0 (0.0) | 0 (0.0) | 3 (6.4) |
| White | 110 (33.4) | 0 (0.0) | 25 (44.6) | 29 (80.6) | 21 (70.0) | 10 (83.3) | 22 (46.8) |
| Latin American | 31 ( 9.4) | 0 (0.0) | 20 (35.7) | 0 (0.0) | 4 (13.3) | 0 (0.0) | 5 (10.6) |
| Black | 6 ( 1.8) | 0 (0.0) | 1 (1.8) | 2 (5.6) | 3 (10.0) | 0 (0.0) | 0 (0.0) |
| Arab | 1 ( 0.3) | 0 (0.0) | 0 (0.0) | 0 (0.0) | 0 (0.0) | 0 (0.0) | 1 (2.1) |
| West Asian | 1 ( 0.3) | 0 (0.0) | 0 (0.0) | 0 (0.0) | 0 (0.0) | 0 (0.0) | 1 (2.1) |
| Admixed | 12 ( 3.6) | 0 (0.0) | 9 (16.1) | 1 (2.8) | 1 (3.3) | 1 (8.3) | 0 (0.0) |
| Unknown | 26 ( 7.9) | 1 (0.7) | 1 (1.8) | 2 (5.6) | 1 (3.3) | 1 (8.3) | 15 (31.9) |

**Table S6.**
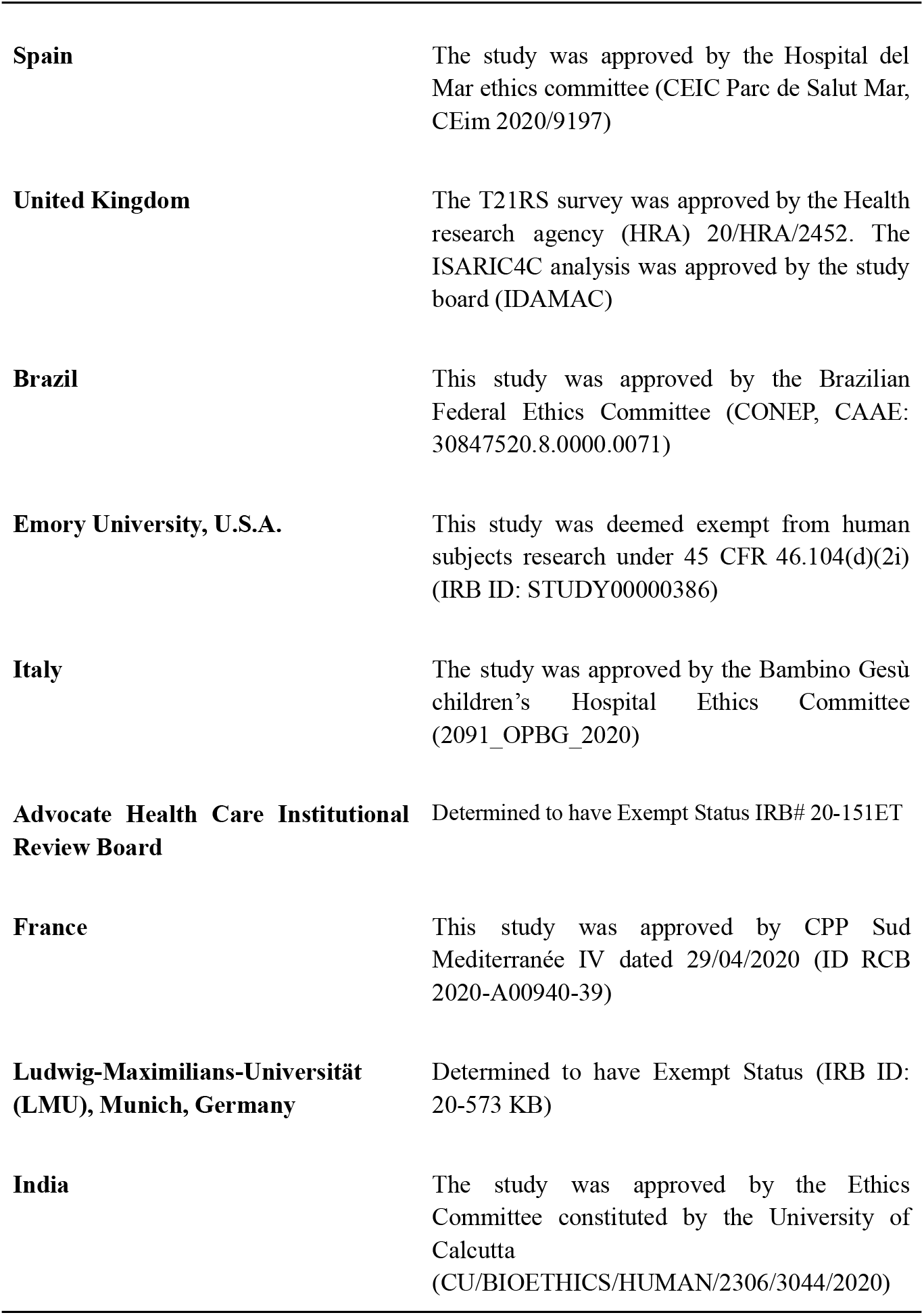
Ethics Approval and Data Protection

**Table S7.**
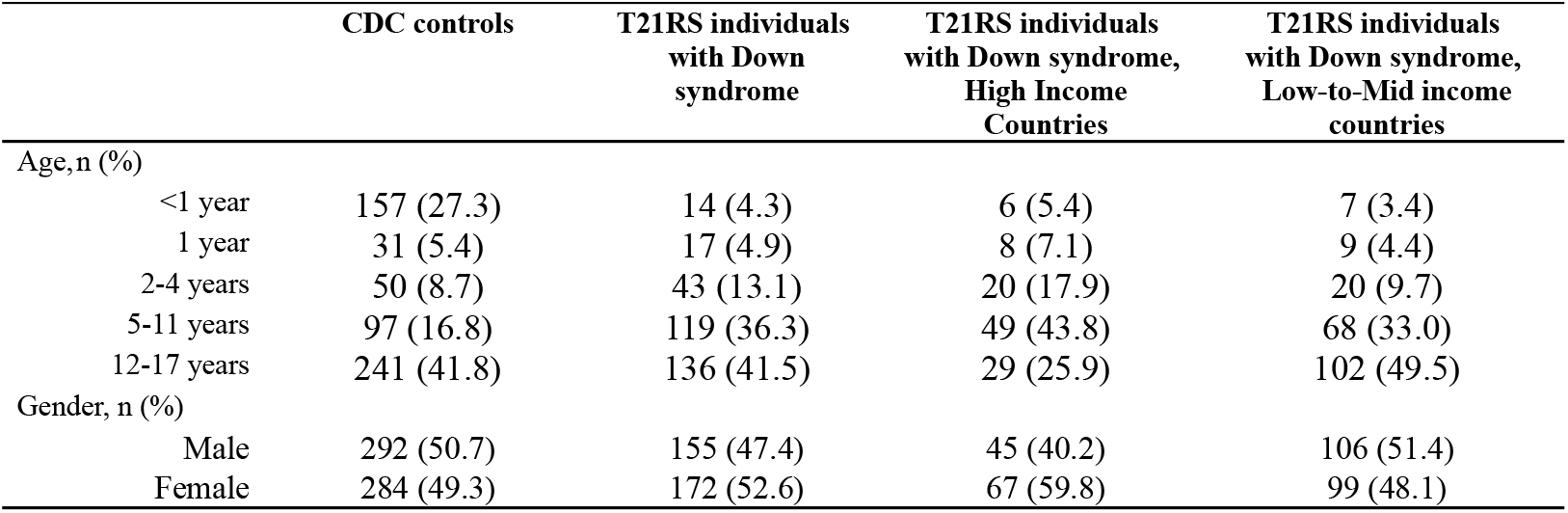
Study characteristics of T21RS individuals with Down syndrome (stratified by High Income and Low-to-Middle Income Countries) and CDC controls without Down syndrome.

|  | CDC controls | T21RS individuals<br>with Down<br>syndrome | T21RS individuals<br>with Down syndrome,<br>High Income<br>Countries | T21RS individuals<br>with Down syndrome,<br>Low-to-Mid income<br>countries |
| --- | --- | --- | --- | --- |
| Age, n (%) |  |  |  |  |
| <1 year | 157 (27.3) | 14 (4.3) | 6 (5.4) | 7 (3.4) |
| 1 year | 31 (5.4) | 17 (4.9) | 8 (7.1) | 9 (4.4) |
| 2-4 years | 50 (8.7) | 43 (13.1) | 20 (17.9) | 20 (9.7) |
| 5-11 years | 97 (16.8) | 119 (36.3) | 49 (43.8) | 68 (33.0) |
| 12-17 years | 241 (41.8) | 136 (41.5) | 29 (25.9) | 102 (49.5) |
| Gender, n (%) |  |  |  |  |
| Male | 292 (50.7) | 155 (47.4) | 45 (40.2) | 106 (51.4) |
| Female | 284 (49.3) | 172 (52.6) | 67 (59.8) | 99 (48.1) |

## Notes

### Competing Interest Statement

The authors have declared no competing interest.

### Clinical Trial

No trial ID as this manuscript uses survey data

### Funding Statement

No external funding was received

### Author Declarations

Spain The study was approved by the Hospital del Mar ethics committee (CEIC Parc de Salut Mar, CEim 2020/9197) United Kingdom The T21RS survey was approved by the Health research agency (HRA) 20/HRA/2452. The ISARIC4C analysis was approved by the study board (IDAMAC) Brazil This study was approved by the Brazilian Federal Ethics Committee (CONEP, CAAE: 30847520.8.0000.0071) Emory University, U.S.A. This study was deemed exempt from human subjects research under 45 CFR 46.104(d)(2i) (IRB ID: STUDY00000386) Italy The study was approved by the Bambino Gesu childrens Hospital Ethics Committee (2091_OPBG_2020) Advocate Health Care Institutional Review Board Determined to have Exempt Status IRB# 20-151ET France This study was approved by CPP Sud Mediterranee IV dated 29/04/2020 (ID RCB 2020-A00940-39) Ludwig-Maximilians-Universitaet (LMU), Munich, Germany Determined to have Exempt Status (IRB ID: 20-573 KB) India The study was approved by the Ethics Committee constituted by the University of Calcutta (CU/BIOETHICS/HUMAN/2306/3044/2020)

## Sources

1. Espinosa J. Down Syndrome and COVID-19: A Perfect Storm?. Cell Rep Med. 2020;1(2):100019. doi:10.1016/j.xcrm.2020.100019

2. Hüls A, Costa A, Dierssen M, Baksh A, Bargagna S, Baumer N. An International Survey on the Impact of COVID-19 in Individuals with Down Syndrome. MedXRiv (Preprint). 2020.

3. De Toma I, Dierssen M. Network analysis of Down syndrome and SARS-CoV-2 identifies risk and protective factors for COVID-19. Sci Rep. 2021;11(1). doi:10.1038/s41598-021-81451-w

4. Prayle A, Vyas H. Respiratory Complications of Down Syndrome. In: Willmott R, ed. Kendig’S Disorders Of The Respiratory Tract In Children. 9th ed. Elsevier; 2019:1161–1202.

5. Carsetti R, Valentini D, Marcellini V et al. Reduced numbers of switched memory B cells with high terminal differentiation potential in Down syndrome. Eur J Immunol. 2014;45(3):903–914. doi:10.1002/eji.201445049

6. Bloemers B, Broers C, Bont L, Weijerman M, Gemke R, van Furth A. Increased risk of respiratory tract infections in children with Down syndrome: the consequence of an altered immune system. Microbes Infect. 2010;12(11):799–808. doi:10.1016/j.micinf.2010.05.007

7. Kantar A, Mazza A, Bonanomi E et al. COVID-19 and children with Down syndrome: is there any real reason to worry? Two case reports with severe course. BMC Pediatr. 2020;20(1). doi:10.1186/s12887-020-02471-5

8. Newman A, Jhaveri R, Patel A, Tan T, Toia J, Arshad M. Trisomy 21 and Coronavirus Disease 2019 in Pediatric Patients. J Pediatr. 2021;228:294–296. doi:10.1016/j.jpeds.2020.08.067

9. Stewart R, El-Harakeh A, Cherian S. Evidence synthesis communities in low-income and middle-income countries and the COVID-19 response. The Lancet. 2020;396(10262):1539–1541. doi:10.1016/s0140-6736(20)32141-3

10. Kim L, Whitaker M, O’Halloran A et al. Hospitalization Rates and Characteristics of Children Aged <18 Years Hospitalized with Laboratory-Confirmed COVID-19 — COVID-NET, 14 States, March 1–July 25, 2020. MMWR Morb Mortal Wkly Rep. 2020;69(32):1081–1088. doi:10.15585/mmwr.mm6932e3

11. High Income Countries 2021. Worldpopulationreview.com. https://worldpopulationreview.com/country-rankings/high-income-countries. Published 2021. Accessed June 8, 2021.

12. Patel N. Pediatric COVID-19: Systematic review of the literature. Am J Otolaryngol. 2020;41(5):102573. doi:10.1016/j.amjoto.2020.102573

13. Costa-Carvalho B, Martinez R, Dias A et al. Antibody response to pneumococcal capsular polysaccharide vaccine in Down syndrome patients. Brazilian Journal of Medical and Biological Research. 2006;39(12):1587–1592. doi:10.1590/s0100-879x2006001200010

14. da Rosa Utiyama S, Nisihara R, Nass F, Oliveira N, Fiedler P, de Messias-Reason I. Autoantibodies in patients with Down Syndrome: Early senescence of the immune system or precocious markers for immunological diseases?. J Paediatr Child Health. 2008;44(4):182–186. doi:10.1111/j.1440-1754.2007.01229.x

15. Valentini D, Marcellini V, Bianchi S et al. Generation of switched memory B cells in response to vaccination in Down syndrome children and their siblings. Vaccine. 2015;33(48):6689–6696. doi:10.1016/j.vaccine.2015.10.083

16. Eijsvoogel N, Hollegien M, Bok V et al. Declining antibody levels after hepatitis B vaccination in Down syndrome: A need for booster vaccination?. J Med Virol. 2017;89(9):1682–1685. doi:10.1002/jmv.24813

17. Palaiodimos L, Kokkinidis D, Li W et al. Severe obesity, increasing age and male sex are independently associated with worse in-hospital outcomes, and higher in-hospital mortality, in a cohort of patients with COVID-19 in the Bronx, New York. Metabolism. 2020;108:154262. doi:10.1016/j.metabol.2020.154262

18. George P, Wells A, Jenkins R. Pulmonary fibrosis and COVID-19: the potential role for antifibrotic therapy. The Lancet Respiratory Medicine. 2020;8(8):807–815. doi:10.1016/s2213-2600(20)30225-3

19. Peckham H, de Gruijter N, Raine C et al. Male sex identified by global COVID-19 meta-analysis as a risk factor for death and ITU admission. Nat Commun. 2020;11(1). doi:10.1038/s41467-020-19741-6

20. French J, Brodie M, Caraballo R et al. Keeping people with epilepsy safe during the COVID-19 pandemic. Neurology. 2020;9(94):1032–1037. doi:https://doi.org/10.1212/wnl.0000000000009632

21. Kuroda N. Epilepsy and COVID-19: Associations and important considerations. Epilepsy & Behavior. 2020;108:107122. doi:10.1016/j.yebeh.2020.107122

22. Asadi-Pooya A, Attar A, Moghadami M, Karimzadeh I. Management of COVID-19 in people with epilepsy: drug considerations. Neurological Sciences. 2020;41(8):2005–2011. doi:10.1007/s10072-020-04549-5

23. Niazkar H, Zibaee B, Nasimi A, Bahri N. The neurological manifestations of COVID-19: a review article. Neurological Sciences. 2020;41(7):1667–1671. doi:10.1007/s10072-020-04486-3

24. Chen M, Zhou W, Xu W. Thyroid Function Analysis in 50 Patients with COVID-19: A Retrospective Study. Thyroid. 2021;31(1):8–11. doi:10.1089/thy.2020.0363

25. Millet J, Whittaker G. Host cell proteases: Critical determinants of coronavirus tropism and pathogenesis. Virus Res. 2015;202:120–134. doi:10.1016/j.virusres.2014.11.021

26. Lazartigues E, Qadir M, Mauvais-Jarvis F. Endocrine Significance of SARS-CoV-2’s Reliance on ACE2. Endocrinology. 2020;161(9). doi:10.1210/endocr/bqaa108

